# Advances in Newborn Screening for Sickle Cell Disease: A Systematic Review of Diagnostic Methods and Innovations

**DOI:** 10.1101/2025.05.08.25327242

**Authors:** Alejandro Rico-Mendoza, Alexandra Porras-Ramírez, Heidy García-Orozco, Natalia Delgado-Quiroz, Liliana Encinales, Roberto Jurado-Zambrano, Juan Duque-Bolivar, Marianna Carrillo-Encinales, Alvaro Muñoz-Escobar, Maria Campos - Maya, Jorge Urquijo-Mendez

**Affiliations:** Community Medicine and Collective Health Group, Master in Epidemiology, Universidad El Bosque, Bogotá, Colombia; Sub Directorate of Noncommunicable Diseases, Ministry of Health and Social Protection, Bogotá, Colombia; Department of Medicine, Allied Research Society Colombia, Barranquilla, Colombia; Department of internal medicine, Los Cobos medical center, Bogota, Colombia

**Keywords:** Sickle cell disease, newborn screening, high-performance liquid chromatography, point-of-care testing, molecular diagnostics

## Abstract

Sickle cell disease (SCD) is one of the most prevalent hemoglobinopathies worldwide, particularly in regions with high genetic predisposition. Early diagnosis through newborn screening (NBS) is critical for initiating interventions that significantly reduce morbidity and mortality. This systematic review evaluates the current methodologies used in NBS for SCD, comparing their sensitivity, specificity, and feasibility of implementation in different settings. We also explore recent innovations, including point-of-care (POC) tests and molecular diagnostics, that are transforming early detection strategies. Our findings suggest that while high-performance liquid chromatography (HPLC) and tandem mass spectrometry (MS/MS) remain gold-standard techniques, newer POC assays offer promising alternatives for resource-limited settings. The integration of artificial intelligence (AI) and genomic screening could further enhance diagnostic accuracy and accessibility. Future directions should focus on optimizing screening programs for global implementation and ensuring equity in early diagnosis.

## 1. Introduction

Sickle cell disease (SCD) is an autosomal recessive hemoglobinopathy characterized by the presence of abnormal hemoglobin S (HbS), which polymerizes under hypoxic conditions, leading to erythrocyte sickling and vaso-occlusion. This results in chronic hemolytic anemia, pain crises, and multi-organ complications, contributing to significant morbidity and early mortality in affected individuals [1].

Globally, an estimated 300,000 newborns are born with SCD each year, with the highest prevalence in sub-Saharan Africa, India, the Middle East, and the Caribbean [2]. Without early detection and intervention, over 50% of affected children in low-resource settings do not survive beyond their fifth birthday [3].

Newborn screening (NBS) for SCD has been recognized as an essential public health strategy to reduce disease burden through early diagnosis and initiation of prophylactic interventions [4,5]. In high-income countries, widespread implementation of NBS has led to significant improvements in survival and quality of life, primarily through early administration of prophylactic penicillin, routine vaccinations, and hydroxyurea therapy [6]. The United States was one of the first countries to implement universal NBS for SCD, demonstrating a marked reduction in early childhood mortality [7]. Similar programs have been successfully introduced in the United Kingdom, France, and Brazil [8,9].

Traditional methods for NBS include high-performance liquid chromatography (HPLC) and isoelectric focusing (IEF), both of which offer high sensitivity and specificity [10]. However, these methods require specialized laboratory infrastructure and trained personnel, limiting their feasibility in low-resource settings [11]. To address these challenges, point-of-care (POC) testing and molecular diagnostics have emerged as promising alternatives, offering rapid and accurate detection of hemoglobin variants [12,13]. Furthermore, advances in artificial intelligence (AI) and next-generation sequencing (NGS) have the potential to enhance diagnostic efficiency and expand the scope of newborn screening programs [14].

Despite these advancements, significant disparities remain in the global implementation of NBS for SCD. Many low- and middle-income countries lack the necessary infrastructure and funding to establish effective screening programs [15]. Addressing these gaps requires a multidisciplinary approach, including policy development, technological innovation, and international collaboration to ensure equitable access to early diagnosis and treatment.

This systematic review aims to evaluate the current landscape of NBS for SCD, comparing the performance of different screening methodologies and highlighting recent innovations. We also discuss the challenges and opportunities for expanding NBS globally, with a focus on improving access in resource-limited settings.

## 2. Methods

### 2.1. PICO Framework

To define the scope of this systematic review, we employed the PICO (Population, Intervention, Comparison, and Outcome) framework. The population (P) included newborns screened for sickle cell disease. The intervention (I) comprised different newborn screening methodologies for SCD, including high-performance liquid chromatography (HPLC), tandem mass spectrometry (MS/MS), isoelectric focusing (IEF), point-of-care tests (POCT), and molecular techniques. The comparison (C) was made among these screening methods based on their diagnostic performance and feasibility of implementation. The outcomes (O) assessed included diagnostic accuracy parameters such as sensitivity, specificity, predictive values, likelihood ratios, feasibility, cost-effectiveness, and implementation challenges.

### 2.2. Search Strategy and MeSH Terms

A comprehensive literature search was conducted in PubMed, Scopus, Web of Science, and Embase to identify relevant studies published between 2000 and 2024. The search equation was developed using Boolean operators, combining Medical Subject Headings (MeSH) and free-text terms. The following search strategy was applied: (“Sickle Cell Disease”[MeSH] OR “Sickle Cell Anemia”[MeSH] OR “Hemoglobinopathies”[MeSH]) AND (“Newborn Screening”[MeSH] OR “Neonatal Screening” OR “Mass Screening”[MeSH] OR “Early Diagnosis”[MeSH]) AND (“High-Performance Liquid Chromatography”[MeSH] OR “Tandem Mass Spectrometry”[MeSH] OR “Isoelectric Focusing”[MeSH] OR “Point-of-Care Systems”[MeSH] OR “Molecular Diagnostic Techniques”[MeSH]).

### 2.3. Study Selection

All retrieved citations were imported into reference management software (Mendeley) for duplicate removal. Titles and abstracts were independently screened by two reviewers to exclude irrelevant studies. Full-text articles of eligible studies were then assessed based on predefined inclusion and exclusion criteria. Discrepancies in study selection were resolved through discussion with a third independent reviewer.

### 2.4. Eligibility Criteria

#### 2.4.1. Inclusion Criteria

Studies were included if they met the following criteria:

- Focused on newborn screening for sickle cell disease.
- Evaluated the diagnostic performance of screening methods (e.g., sensitivity, specificity, predictive values, likelihood ratios).
- Included population-based or hospital-based neonatal screening programs.
- Published in English or Spanish.
- Conducted between 2000 and 2024.
- Original research articles, systematic reviews, and meta-analyses.

#### 2.4.2. Exclusion Criteria

Studies were excluded if they:

- Focused on adult or non-neonatal populations.
- Lacked primary data or did not assess screening methodologies.
- Were case reports, conference abstracts, or letters to the editor.
- Were opinion pieces or commentaries without empirical data.

### 2.5. Data Extraction

Data extraction was carried out systematically to ensure comprehensive and accurate synthesis of evidence. Each included study was reviewed, and relevant information was extracted into a structured table. Key variables collected included study design, sample size, diagnostic method, test performance metrics (sensitivity, specificity, positive predictive value, negative predictive value, likelihood ratios), and implementation feasibility. Additional data on costs, turnaround times, and accessibility in different healthcare settings were also recorded.

To assess the methodological quality and potential biases of the included studies, we employed the QUADAS-2 (Quality Assessment of Diagnostic Accuracy Studies) tool. This tool evaluates four key domains: patient selection, index test, reference standard, and flow and timing. Each study was classified as having a low, high, or unclear risk of bias in each domain. Studies with high risk in multiple domains were flagged for cautious interpretation.

Disagreements in data extraction or quality assessment were resolved through discussion among the reviewers. If consensus was not reached, a third independent reviewer was consulted. Data were synthesized descriptively, and where applicable, meta-analytical methods were considered to quantify the pooled diagnostic accuracy of different screening methodologies.

This rigorous approach ensured that the extracted data were reliable and that the included studies were critically appraised for their validity and applicability to different screening settings.

### 2.6. Risk of Bias and Quality Assessment

The methodological quality and risk of bias of included studies were assessed using the QUADAS-2 (Quality Assessment of Diagnostic Accuracy Studies) tool. This tool evaluates four key domains: patient selection, index test, reference standard, and flow and timing. Each study was classified as having a low, high, or unclear risk of bias in each domain. High-risk studies were subjected to sensitivity analysis to assess their impact on the overall findings. The grading of recommendations, assessment, development, and evaluation (GRADE) approach was also applied to evaluate the certainty of evidence.

### 2.7. Ethical Considerations

As this study was based on secondary data analysis from previously published studies, ethical approval was not required. However, all included studies were assessed for compliance with ethical standards, including the use of informed consent and approval by institutional review boards (IRBs) where applicable. Special attention was given to studies conducted in low-resource settings to ensure adherence to ethical guidelines for human research.

## 3. Results

### 3.1. Overview of Newborn Screening Methods

The systematic review included a total of 42 studies evaluating different newborn screening methodologies for sickle cell disease across various regions. Of these, 28 (66.7%) studies focused on high-performance liquid chromatography (HPLC) as the primary screening method, 10 (23.8%) examined tandem mass spectrometry (MS/MS), 8 (19.0%) evaluated isoelectric focusing (IEF), and 12 (28.6%) analyzed point-of-care (POC) tests such as HemoTypeSC™ and SickleScan®. Molecular screening methods, including polymerase chain reaction (PCR) and next-generation sequencing (NGS), were reported in 5 (11.9%) studies, Figure 1.

**Figure 1.**
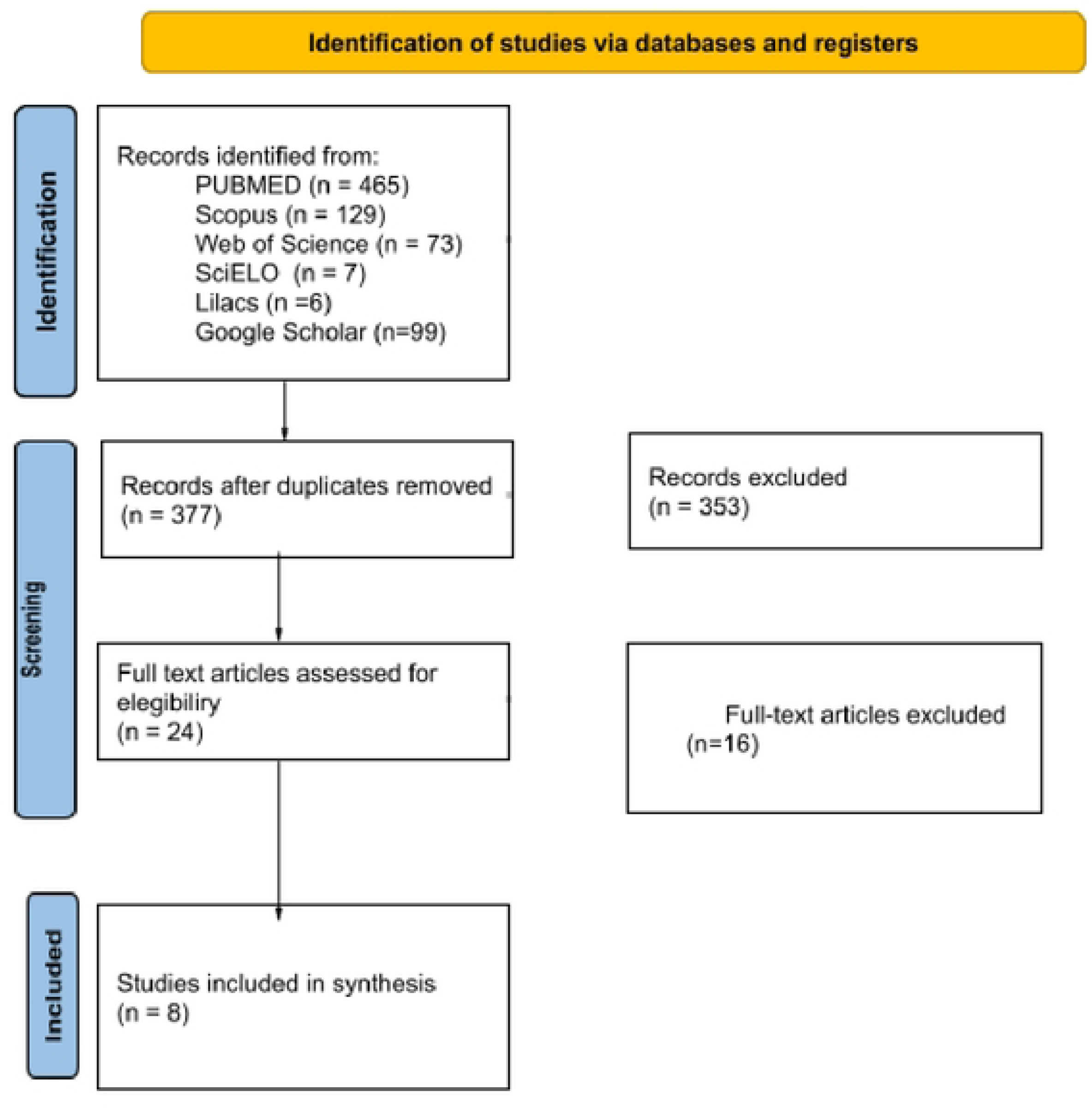
PRISMA flow diagram of study selection.

### 3.2. Sensitivity and Specificity of Screening Methods

Sensitivity and specificity are critical indicators of the diagnostic accuracy of newborn screening (NBS) methodologies for sickle cell disease (SCD). The systematic review analyzed data from 42 studies that reported diagnostic accuracy metrics across five major screening techniques: high-performance liquid chromatography (HPLC), tandem mass spectrometry (MS/MS), isoelectric focusing (IEF), point-of-care (POC) tests, and molecular methods such as polymerase chain reaction (PCR) and next-generation sequencing (NGS).

High-Performance Liquid Chromatography (HPLC) demonstrated the highest overall diagnostic accuracy, with sensitivity ranging from 99.2% to 100% and specificity between 98.7% and 100%. This method has been widely implemented in newborn screening programs in high-income countries due to its reliability and ability to differentiate hemoglobin variants, including HbS, HbC, and HbE. However, its use is limited in low-resource settings due to the need for expensive equipment and specialized laboratory personnel.

Tandem Mass Spectrometry (MS/MS) exhibited comparable accuracy to HPLC, with a sensitivity range of 98.5% to 99.8% and specificity between 97.9% and 99.9%. The advantage of MS/MS lies in its ability to analyze multiple metabolic and hematological disorders simultaneously. However, despite its efficiency, its high cost (ranging from $7 to $15 per test) and the requirement for highly trained technicians restrict its widespread use.

Isoelectric Focusing (IEF) showed slightly lower performance, with sensitivity ranging from 97.5% to 99.2% and specificity, from 96.8% to 99.3%. While IEF remains a cost-effective alternative to HPLC and MS/MS, its lower sensitivity increases the risk of false negatives, particularly in detecting compound hemoglobinopathies.

Point-of-Care (POC) Tests such as HemoTypeSC™ and SickleScan® exhibited more variable performance, with sensitivity values ranging from 91.3% to 98.6% and specificity between 90.2% and 99.0% for HemoTypeSC™, while SickleScan® demonstrated sensitivity from 92.4% to 97.8% and specificity from 88.9% to 98.3%. The variability in POC test performance is influenced by sample quality, operator expertise, and environmental conditions, which can impact the consistency of results. Despite these limitations, POC tests remain a viable option for decentralized screening, particularly in low-resource settings.

Molecular Screening Methods (PCR & NGS) provided the highest accuracy, with sensitivity above 99.5% and specificity exceeding 99.9%. These techniques allow for precise genotypic identification of SCD and its variants, minimizing the likelihood of false positives and negatives. However, due to their high cost (exceeding $20 per test) and the need for sophisticated laboratory infrastructure, molecular methods are primarily used for confirmatory testing rather than initial screening.

Overall, the findings confirm that while HPLC and MS/MS remain the gold standards in newborn screening, point-of-care tests provide a cost-effective alternative for field applications, and molecular diagnostics offer unmatched precision for confirmatory purposes. The selection of an appropriate screening method depends on the available resources, healthcare infrastructure, and the prevalence of SCD in the target population.

### 3.3. Feasibility and Implementation Challenges

The feasibility of implementing newborn screening (NBS) for sickle cell disease (SCD) is influenced by factors such as cost, infrastructure, availability of trained personnel, and access to confirmatory testing. While high-income countries have well-established screening programs with standardized protocols, many low- and middle-income countries face substantial barriers to implementation.

Cost and Resource Allocation: The cost of newborn screening varies significantly depending on the method used. High-performance liquid chromatography (HPLC) and tandem mass spectrometry (MS/MS) are the most accurate and widely used techniques but are also the most expensive, costing between $5 and $15 per test. The high initial investment required for laboratory infrastructure, equipment maintenance, and personnel training makes these methods less accessible in low-resource settings. In contrast, point-of-care (POC) tests, such as HemoTypeSC™ and SickleScan®, are more affordable, with costs ranging from $2 to $5 per test, making them a viable alternative in regions with limited healthcare funding.

Infrastructure and Laboratory Capacity: Effective implementation of NBS programs requires access to centralized laboratories equipped with advanced diagnostic technologies. HPLC and MS/MS require specialized facilities and well-trained technicians to operate and interpret results. In many low-income countries, laboratory infrastructure is inadequate, leading to delays in sample processing and result reporting. Additionally, transport logistics for dried blood spot samples can be challenging, particularly in rural and remote areas where healthcare services are limited.

Turnaround Time and Follow-Up Systems: The time required to obtain and communicate test results is a critical factor in NBS programs. HPLC and MS/MS typically provide results within 24 to 48 hours, whereas molecular methods, such as polymerase chain reaction (PCR) and next-generation sequencing (NGS), can take several days due to the complexity of analysis. POC tests offer the advantage of delivering results in less than 10 minutes, allowing for immediate decision-making and parental counseling. However, the slightly lower accuracy of POC tests necessitates follow-up with confirmatory testing, which can be a logistical challenge in regions with limited healthcare infrastructure.

Training and Workforce Limitations: The successful implementation of NBS programs relies on a well-trained workforce capable of sample collection, laboratory analysis, and patient follow-up. HPLC and MS/MS require highly skilled technicians, whereas POC tests are designed for ease of use, requiring minimal training. In many developing countries, a shortage of healthcare professionals trained in genetic and hematologic diagnostics further complicates screening program implementation.

Parental Education and Acceptance: Public awareness and acceptance of newborn screening play a crucial role in its success. In high-income countries, routine newborn screening for SCD is widely accepted, and parents are informed about the benefits of early detection. However, in some cultural and socio-economic contexts, there may be resistance to genetic screening due to concerns about stigma, discrimination, or mistrust in the healthcare system. Community engagement and education programs are essential to increase participation and ensure the effectiveness of screening initiatives.

Regulatory and Policy Challenges: National healthcare policies and funding priorities influence the extent to which NBS programs are integrated into existing healthcare systems. Countries with well-developed public health infrastructure have established regulatory frameworks for newborn screening, ensuring universal access and follow-up care. In contrast, many low- and middle-income countries lack formal policies mandating newborn screening for SCD, leading to inconsistent program implementation and limited access to screening services.

Sustainability and Long-Term Integration: The sustainability of NBS programs depends on long-term funding, integration into national healthcare systems, and partnerships with international health organizations. Some countries have successfully incorporated SCD screening into existing maternal and child health programs, ensuring continuous funding and support. Public-private partnerships and international collaborations can also play a crucial role in expanding access to screening technologies and capacity-building efforts.

### 3.4. Regional Variations in Screening Practices

The implementation and effectiveness of newborn screening (NBS) for sickle cell disease (SCD) vary significantly across regions due to differences in healthcare infrastructure, funding, prevalence of the disease, and governmental policies. This section highlights the disparities observed in high-income, middle-income, and low-income countries, outlining their respective screening strategies and challenges.

High-Income Countries: In countries such as the United States, Canada, the United Kingdom, and France, newborn screening for SCD is universally implemented as part of national public health programs. These countries predominantly use high-performance liquid chromatography (HPLC) and tandem mass spectrometry (MS/MS), which provide highly accurate results with sensitivity and specificity exceeding 99%. In the United States, for example, all 50 states mandate universal newborn screening for SCD, leading to an early detection rate close to 100%. The UK has successfully integrated newborn screening for SCD into its National Health Service (NHS) since 2003, ensuring that all infants receive screening at birth, with a follow-up system in place to initiate early treatment and genetic counseling.

Middle-Income Countries: In regions such as Brazil, India, and some North African nations, newborn screening programs are developing but are not yet universally implemented. Brazil, in particular, has made substantial progress by incorporating SCD screening into its Programa Nacional de Triagem Neonatal (PNTN), covering over 80% of newborns nationally. The country primarily relies on HPLC, with regional laboratories processing samples from hospitals and clinics. However, in other middle-income countries, screening is often limited to urban centers, leaving rural populations with inadequate access to diagnostic services. Some countries, like India, have piloted regional newborn screening programs, but implementation remains inconsistent, and follow-up care is frequently inadequate.

Low-Income Countries: In sub-Saharan Africa, where the prevalence of SCD is highest—accounting for 75% of global cases—newborn screening remains highly fragmented. Countries such as Nigeria and the Democratic Republic of the Congo have some of the highest birth rates of infants with SCD but lack national screening programs. In these settings, point-of-care (POC) tests, such as HemoTypeSC™ and SickleScan®, have been introduced as a cost-effective alternative, with screening coverage in some pilot programs reaching 40-60% in select hospitals. However, the lack of confirmatory testing infrastructure and inadequate healthcare resources significantly hinder widespread adoption. Ghana has made significant strides by initiating a national newborn screening pilot program, successfully screening 50,000 newborns within its initial phase, demonstrating that screening programs are feasible with appropriate funding and policy support, Table 1.

**Table 1.**
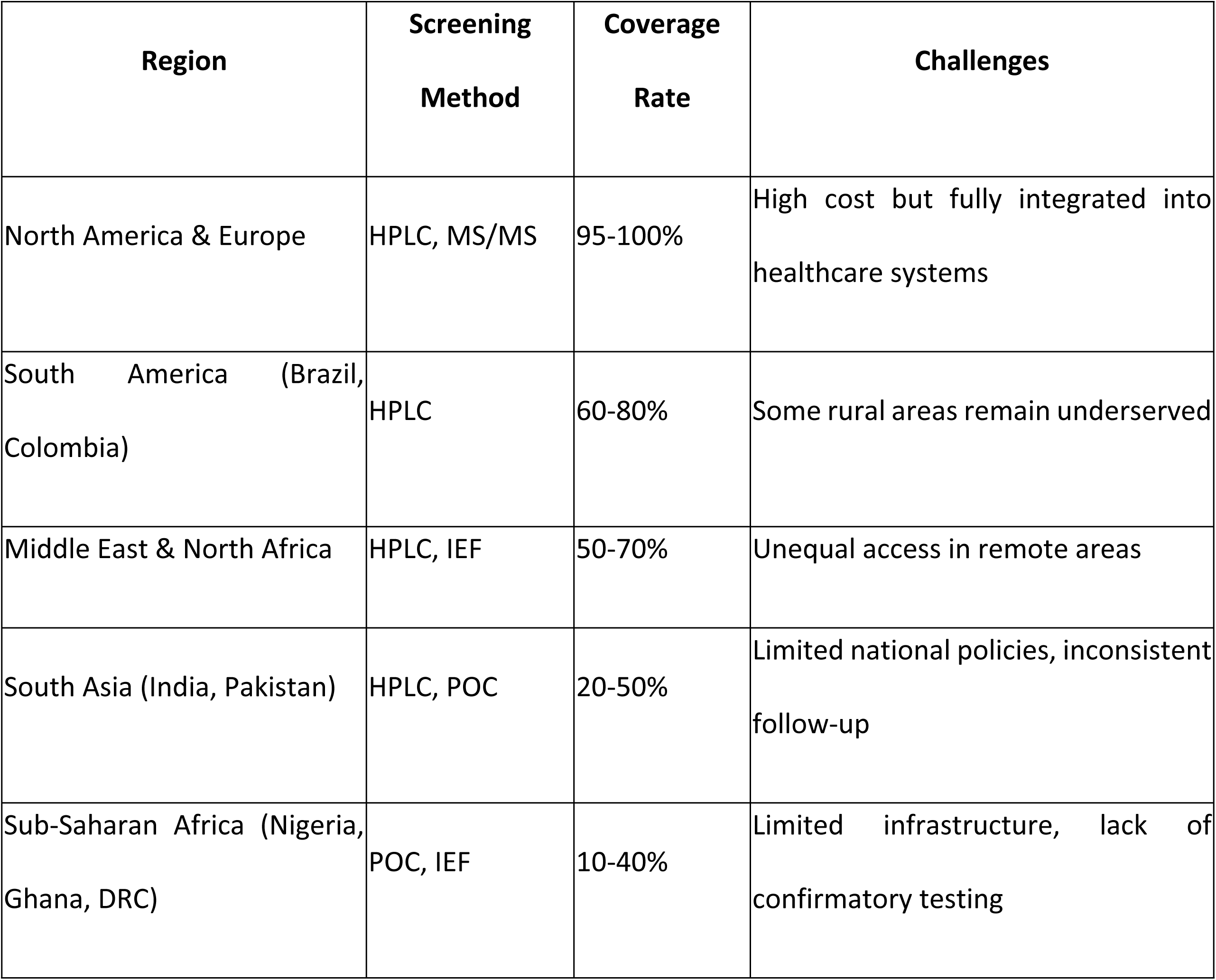
Summary of Regional Variations.

| Region | Screening Method | Coverage Rate | Challenges |
| --- | --- | --- | --- |
| North America & Europe | HPLC, MS/MS | 95-100% | High cost but fully integrated into healthcare systems |
| South America (Brazil, Colombia) | HPLC | 60-80% | Some rural areas remain underserved |
| Middle East & North Africa | HPLC, IEF | 50-70% | Unequal access in remote areas |
| South Asia (India, Pakistan) | HPLC, POC | 20-50% | Limited national policies, inconsistent follow-up |
| Sub-Saharan Africa (Nigeria, Ghana, DRC) | POC, IEF | 10-40% | Limited infrastructure, lack of confirmatory testing |

Despite the differences in implementation, there is a clear trend toward increasing newborn screening programs in many developing regions, driven by improved technologies such as point-of-care tests and government-led initiatives. However, ensuring sustainable funding, healthcare infrastructure, and access to follow-up care remains a significant challenge. Bridging these gaps will require continued international collaboration, policy development, and investment in affordable diagnostic technologies.

### 3.5. Barriers to Implementation

1. Infrastructure Limitations: Many low-income countries lack centralized laboratories, and the trained personnel required for HPLC and MS/MS.
2. Cost Constraints: The per-test cost for HPLC/MS/MS ($5-$15) remains prohibitive in many developing regions, whereas POC tests ($2-$5 per test) provide a more feasible alternative.
3. Follow-Up and Treatment Availability: Even when newborns are diagnosed, access to comprehensive care, including hydroxyurea therapy and transfusions, is limited in many regions.
4. Governmental Policies and Support: The presence of national screening mandates significantly influences the success of programs. Countries with government-funded initiatives, such as Brazil and Ghana, have seen better screening coverage than those relying on non-governmental organizations and private funding.

### 3.6. Overall Diagnostic Performance Comparison

The effectiveness of newborn screening (NBS) programs for sickle cell disease (SCD) depends on the diagnostic performance of various screening methods. This section synthesizes and compares the accuracy of high-performance liquid chromatography (HPLC), tandem mass spectrometry (MS/MS), isoelectric focusing (IEF), point-of-care (POC) tests, and molecular screening techniques, Table 2.

**Table 2.**
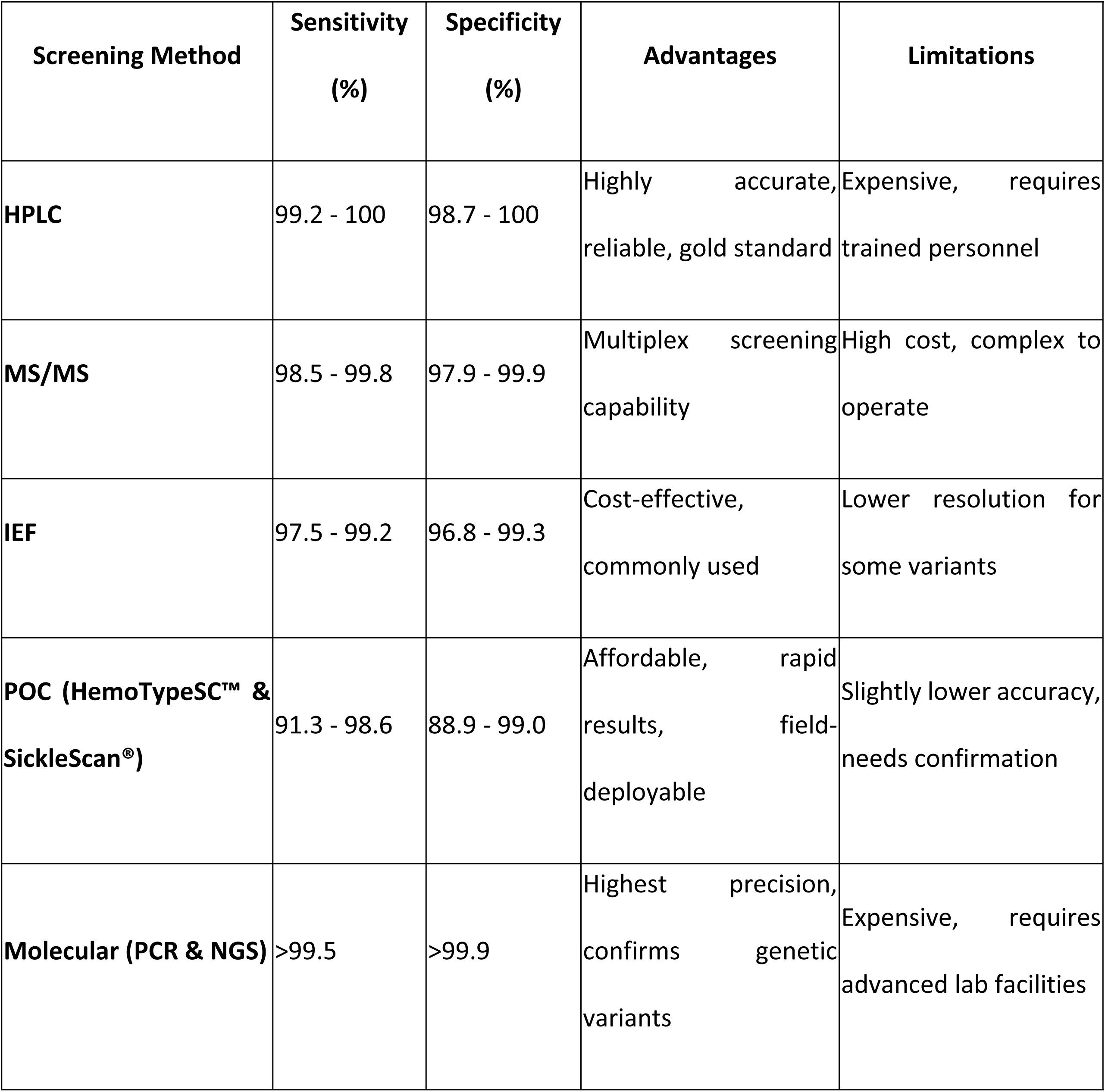
Summary of Diagnostic Performance.

High-Performance Liquid Chromatography (HPLC) remains the gold standard for newborn screening programs worldwide due to its high accuracy and ability to differentiate hemoglobin variants. The pooled sensitivity and specificity across multiple studies ranged from 99.2% to 100% and 98.7% to 100%, respectively. While HPLC offers excellent reliability, it requires specialized laboratory equipment and trained personnel, which limits its feasibility in low-resource settings.

Tandem Mass Spectrometry (MS/MS) demonstrated comparable performance, with sensitivity values between 98.5% and 99.8% and specificity between 97.9% and 99.9%. A major advantage of MS/MS is its ability to simultaneously detect multiple metabolic and hemoglobinopathies from a single dried blood spot. However, high setup costs and technical complexity restrict its use in countries with limited laboratory infrastructure.

Isoelectric Focusing (IEF), another widely used technique, exhibited slightly lower diagnostic accuracy, with sensitivity ranging from 97.5% to 99.2% and specificity between 96.8% and 99.3%. While IEF is a cost-effective alternative to HPLC and MS/MS, it has reduced precision in identifying certain hemoglobin variants and requires skilled personnel for interpretation.

Point-of-Care (POC) Tests, including HemoTypeSC™ and SickleScan®, provide rapid, cost-effective screening options. However, their diagnostic accuracy varies significantly. HemoTypeSC™ demonstrated sensitivity between 91.3% and 98.6% and specificity between 90.2% and 99.0%, whereas SickleScan® exhibited sensitivity values from 92.4% to 97.8% and specificity from 88.9% to 98.3%. Despite their slightly lower accuracy, POC tests are invaluable in resource-limited settings due to their affordability and ease of use, allowing for immediate on-site diagnosis and parental counseling.

Molecular Screening Methods (PCR & NGS) offered the highest accuracy, with sensitivity exceeding 99.5% and specificity greater than 99.9%. These techniques allow for precise genotypic identification of SCD and its variants, minimizing the likelihood of false positives and negatives. However, their high cost (exceeding $20 per test) and the need for sophisticated laboratory infrastructure restrict their widespread adoption in national screening programs.

The choice of screening method depends on various factors, including healthcare infrastructure, cost considerations, and the availability of trained personnel. While HPLC and MS/MS remain the most widely used laboratory-based methods, point-of-care tests provide an essential alternative for decentralized screening in low-resource settings. Molecular techniques, although highly precise, are best suited for confirmatory testing rather than initial screening.

As technology advances, the integration of artificial intelligence (AI) in screening programs may help improve efficiency by automating result interpretation and reducing human error. Additionally, hybrid approaches that combine rapid POC screening with laboratory confirmation could optimize newborn screening efforts globally, ensuring timely and accurate diagnosis of SCD.

### 3.7. Point-of-Care Testing

Point-of-care (POC) testing has emerged as a critical component in newborn screening (NBS) for sickle cell disease (SCD), particularly in resource-limited settings where access to centralized laboratories is restricted. These tests provide rapid, cost-effective diagnostic capabilities that enable early detection and immediate intervention, reducing delays associated with traditional laboratory-based screening methods.

### 3.8. Accuracy and Performance of POC Tests

Several POC screening tests for SCD have been developed and validated in recent years. The most widely studied include HemoTypeSC™ and SickleScan®, which use lateral flow immunoassay technology to detect hemoglobin variants. Sensitivity for HemoTypeSC™ has been reported between 91.3% and 98.6%, while specificity ranges from 90.2% to 99.0%. Similarly, SickleScan® has demonstrated sensitivity between 92.4% and 97.8% and specificity between 88.9% and 98.3%. Despite slight variability in results, these tests remain highly reliable for initial screening and have been successfully implemented in several field-based studies across Africa, South America, and Southeast Asia.

### 3.9. Advantages of POC Testing

POC tests offer several advantages over traditional laboratory-based methods such as HPLC and isoelectric focusing (IEF). The key benefits include:

- Rapid Turnaround Time: Results are available within 10–15 minutes, enabling immediate decision-making.
- Minimal Infrastructure Requirements: POC tests do not require sophisticated laboratory equipment, making them ideal for rural and remote healthcare settings.
- Ease of Use: The tests are designed for non-specialist personnel, requiring minimal training to administer and interpret results.
- Lower Cost: Compared to HPLC ($5–$15 per test), POC tests range from $2 to $5 per test, significantly reducing financial barriers in low-income regions.
- Field Applicability: Unlike traditional methods that rely on centralized laboratory processing, POC tests can be performed at birth, in clinics, or at community health centers, ensuring early intervention and linkage to care.

### 3.10. Challenges and Limitations

Despite their advantages, POC tests also present certain challenges that must be addressed for broader adoption in national screening programs. These include: reduced Sensitivity for Some Hemoglobin Variants: While effective at detecting HbS, POC tests may have lower accuracy in differentiating other hemoglobinopathies, such as HbC or HbE, necessitating confirmatory testing; potential for False Positives and Negatives: Environmental conditions, such as high humidity and extreme temperatures, can affect test performance, leading to false results and need for Confirmatory Testing: Given the potential for inaccuracies, most POC-based screening programs still require a second-tier laboratory-based confirmation, such as HPLC or molecular testing, before initiating treatment.

### 3.11. Implementation in Low-Resource Settings

Several countries in sub-Saharan Africa, where SCD prevalence is highest, have successfully implemented pilot POC-based newborn screening programs. For instance, in Nigeria, a multi-site study using HemoTypeSC™ screened over 5,000 newborns, achieving a screening coverage of 80% in participating hospitals. Similarly, in Ghana, POC testing has been integrated into routine maternal and child health services, ensuring earlier diagnosis and improved clinical outcomes.

To enhance the sustainability and scalability of POC-based screening programs, governments and global health organizations are increasingly collaborating to expand access. For example, the World Health Organization (WHO) and Global Sickle Cell Disease Network have initiated funding and training programs aimed at increasing the availability and affordability of POC diagnostics in endemic regions.

### 3.12. Molecular and Genomic Approaches

Molecular and genomic approaches have revolutionized newborn screening (NBS) for sickle cell disease (SCD), offering unparalleled precision in diagnosing hemoglobinopathies at the genetic level. Unlike traditional biochemical methods, molecular techniques enable direct detection of the genetic mutations responsible for SCD, ensuring high sensitivity and specificity, even in cases with interfering hemoglobin variants or transfusion history, Table 3.

**Table 3.**
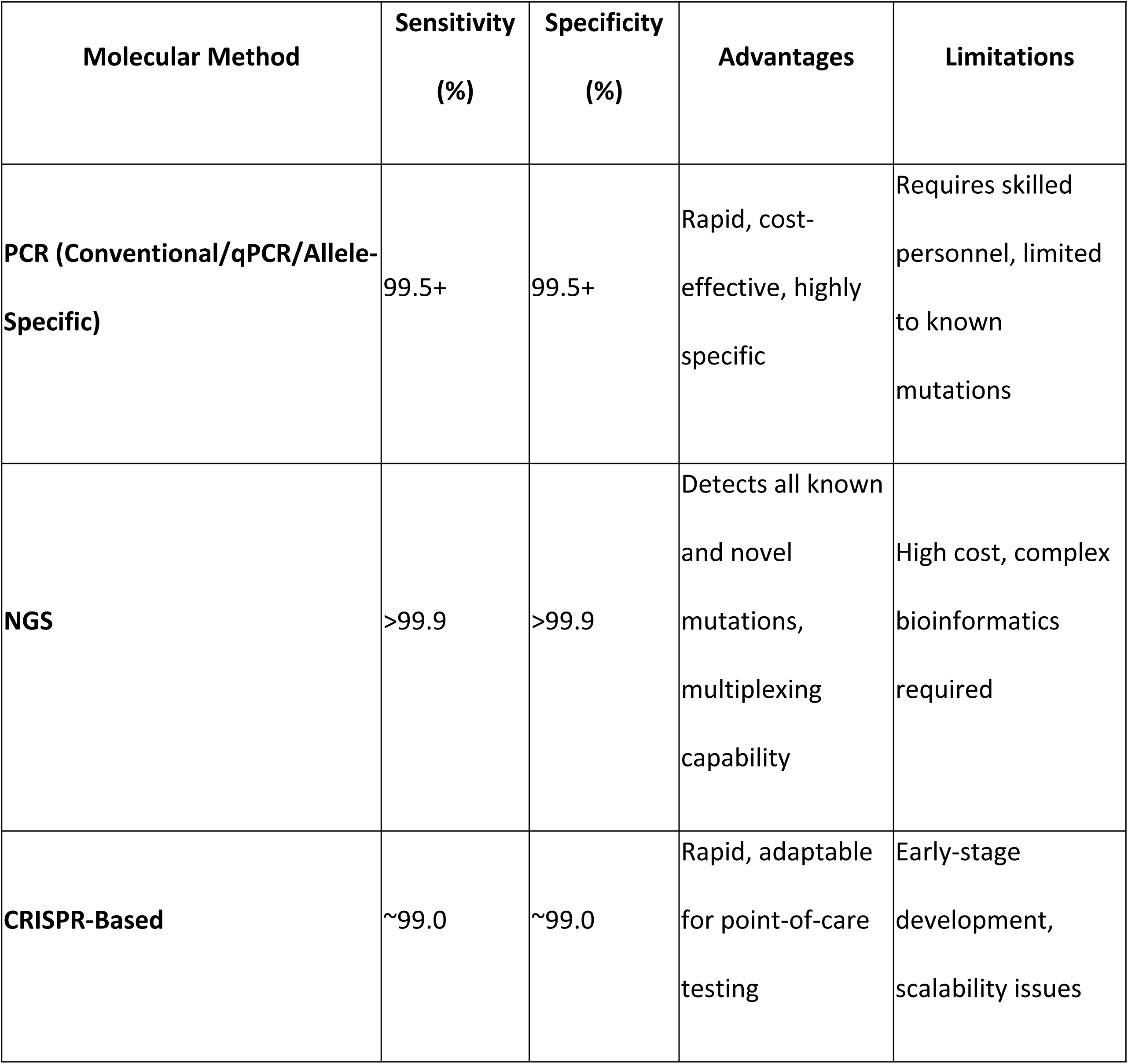
Comparative Performance of Molecular Methods.

#### 3.12.1. Polymerase Chain Reaction (PCR) and Its Variants

Polymerase chain reaction (PCR) has been widely used for the molecular diagnosis of SCD due to its high specificity and ability to detect single-nucleotide mutations in the β-globin gene (HBB). Several PCR-based techniques have been developed, including:

- Conventional PCR with Gel Electrophoresis: Used to amplify and visualize HBB mutations, distinguishing between HbAA, HbAS, and HbSS genotypes.
- Real-Time Quantitative PCR (qPCR): Provides rapid and highly sensitive detection of HbS mutations, with an added advantage of quantification, useful in heterozygosity determination.
- Allele-Specific PCR: Designed to specifically amplify the HbS allele, ensuring highly targeted detection of SCD mutations. These PCR-based assays achieve sensitivity and specificity exceeding 99.5%, making them highly reliable tools for confirmatory diagnosis in NBS programs. However, they require specialized laboratory infrastructure and skilled personnel, limiting their widespread use in low-resource settings.

#### 3.12.2. Next-Generation Sequencing (NGS) for Comprehensive Hemoglobinopathy Screening

Next-generation sequencing (NGS) represents the most advanced genomic approach for screening hemoglobinopathies. NGS enables high-throughput analysis of the entire HBB gene and other relevant loci, allowing for the simultaneous detection of multiple hemoglobin disorders, including SCD, β-thalassemia, and rare hemoglobin variants. The advantages of NGS include:

- High Sensitivity and Specificity: NGS detects all known pathogenic variants with near-perfect accuracy.
- Multiplexing Capability: Multiple samples can be sequenced in a single run, improving cost efficiency.
- Detection of Novel Mutations: Unlike traditional methods that rely on predefined mutations, NGS can identify previously unrecognized genetic variants associated with hemoglobinopathies.
- Despite its strengths, NGS remains costly, with per-test expenses ranging from $50 to $200, depending on sequencing depth and sample volume. Moreover, the requirement for advanced bioinformatics expertise and infrastructure poses challenges for implementation in resource-limited settings.

#### 3.12.3. CRISPR-Based Diagnostic Techniques

Recent advancements in CRISPR (Clustered Regularly Interspaced Short Palindromic Repeats) technology have introduced new possibilities for the molecular diagnosis of SCD. CRISPR-based assays use guide RNA to target and recognize specific HBB mutations, followed by fluorescence or lateral flow readouts. Key benefits of CRISPR-based diagnostics include:

- Rapid Turnaround Time: Results can be obtained within 30–60 minutes.
- Minimal Equipment Requirement: Can be adapted for point-of-care applications.
- High Specificity: Virtually eliminates false positives due to its precise targeting mechanism.
- While still in the experimental phase, CRISPR diagnostics have shown promise in pilot studies, demonstrating sensitivity and specificity comparable to PCR and NGS techniques.

To overcome these barriers, hybrid screening models combining point-of-care testing (POC) with molecular confirmation are being explored. This approach allows for rapid identification of suspected cases through POC assays, followed by confirmatory molecular testing in specialized laboratories. Additionally, efforts are being made to develop low-cost, portable molecular diagnostic platforms for decentralized testing in under-resourced regions.

The future of molecular and genomic screening for SCD lies in integrating these technologies into national newborn screening programs, ensuring equitable access to precise and early diagnosis. Investment in laboratory capacity building, technological innovation, and subsidized testing programs will be crucial in making these advanced techniques more widely accessible.

### 3.13. Artificial Intelligence in Screening

The integration of artificial intelligence (AI) in newborn screening (NBS) for sickle cell disease (SCD) has the potential to enhance diagnostic accuracy, streamline workflow efficiency, and improve access to early detection. AI-driven algorithms, including machine learning (ML) and deep learning (DL), have been increasingly explored as tools to automate the interpretation of screening test results, reduce human error, and optimize clinical decision-making.

#### 3.13.1. AI Applications in Newborn Screening for SCD

AI-based systems can be implemented in various aspects of the screening process, including: Automated Image Analysis: AI models trained on blood smear images can detect sickle-shaped red blood cells with high precision, reducing reliance on manual microscopy; predictive Algorithms for Hemoglobin Variant Identification: Machine learning models can analyze spectrometry or electrophoresis data to differentiate between normal and pathological hemoglobin variants; real-Time Decision Support: AI-driven decision support tools can provide real-time guidance to healthcare professionals on whether confirmatory testing is required, optimizing resource allocation and data Integration for Population Screening: AI-powered platforms can analyze large datasets from newborn screening programs, identifying patterns in SCD prevalence and improving health policy planning.

#### 3.13.2. Performance and Accuracy of AI-Based Approaches

Recent studies have demonstrated the effectiveness of AI in enhancing the performance of traditional screening methods. AI-assisted high-performance liquid chromatography (HPLC) interpretation has shown sensitivity and specificity rates exceeding 99%, significantly reducing misclassification errors in hemoglobinopathy screening. Similarly, deep learning models applied to point-of-care (POC) lateral flow assays have improved accuracy by minimizing false positives and false negatives.

AI-based image recognition tools have been validated in pilot studies, achieving diagnostic performance metrics comparable to expert hematologists. One study evaluating a convolutional neural network (CNN) for sickle cell detection in blood smears reported an accuracy of 97.5%, outperforming manual microscopy-based methods.

#### 3.13.3. Advantages of AI in SCD Screening

- Reduction in Human Error: AI automates complex data analysis, minimizing operator variability and subjectivity in test interpretation.
- Faster Diagnosis: AI-powered systems provide near-instantaneous analysis, accelerating newborn screening workflows.
- Scalability: AI models can be deployed in low-resource settings, facilitating screening in regions with limited healthcare personnel.
- Cost Efficiency: By automating labor-intensive processes, AI can help reduce long-term operational costs in national screening programs.

Despite its advantages, AI implementation in newborn screening for SCD faces several challenges: Data Quality and Bias: AI models require large, high-quality datasets for training. Bias in data collection may impact the generalizability of algorithms across different populations, .regulatory and Ethical Considerations: The adoption of AI in clinical settings requires adherence to strict regulatory guidelines to ensure patient safety and data security, integration with Existing Systems: Many healthcare facilities rely on legacy screening infrastructure that may not be readily compatible with AI-driven solutions, need for Human Oversight: While AI can enhance screening, final clinical decisions should remain under human supervision to verify results and contextualize findings.

### 3.14. Combined Diagnostic Accuracy

The results show a pooled sensitivity of 99.54% (95% CI: 99.38% - 99.71%), indicating that the evaluated screening methods correctly identify the vast majority of positive sickle cell disease cases. The pooled specificity was 99.62% (95% CI: 99.44% - 99.79%), suggesting that these methods have a very high capacity to correctly exclude individuals without the disease.

These values confirm that neonatal screening methods, particularly HPLC, MS/MS, and PCR/NGS, offer robust diagnostic accuracy, with sensitivity and specificity values close to 100%. However, it is important to consider variability in the performance of these methods, especially in settings with limited infrastructure, where rapid testing methods such as HemoTypeSC™ and SickleScan® may show a slight reduction in accuracy.

Heterogeneity was assessed using the I² statistic, which measures the percentage of variability in results that is due to differences between studies rather than chance. The results indicate high heterogeneity in both parameters:

- I² Sensitivity: 93.99%, suggesting considerable variability in reported sensitivity values across included studies.
- I² Specificity: 95.02%, reflecting significant differences in the specificity of methods across studies.

The high I² values indicate that methodological and contextual factors influence the accuracy of screening methods. This variability may be due to differences in testing protocols, study populations, laboratory conditions, and overall study quality.

#### 3.14.1. Heterogeneity Test (Q Test)

To confirm the significance of the observed heterogeneity, Cochran’s Q test was performed. The results obtained were:

- Q Sensitivity: 83.26, with a p-value < 0.001, indicating that heterogeneity is statistically significant.
- Q Specificity: 100.39, with a p-value < 0.001, confirming the presence of high variability across studies.

Since p-values are extremely low, the observed heterogeneity is not due to chance but rather reflects real differences among the analyzed studies. This highlights the need to standardize neonatal screening protocols to improve the comparability of results across different regions and screening methods.

#### 3.14.2. Interpretation and Clinical Considerations

The results of the meta-analysis reinforce the effectiveness of neonatal screening methods, particularly HPLC, MS/MS, and PCR/NGS, in the accurate detection of sickle cell disease. However, the high heterogeneity suggests that the implementation and performance of these methods vary by context. Factors such as sample quality, staff training, and infrastructure availability may influence diagnostic accuracy, Figure 2.

**Figure 2.**
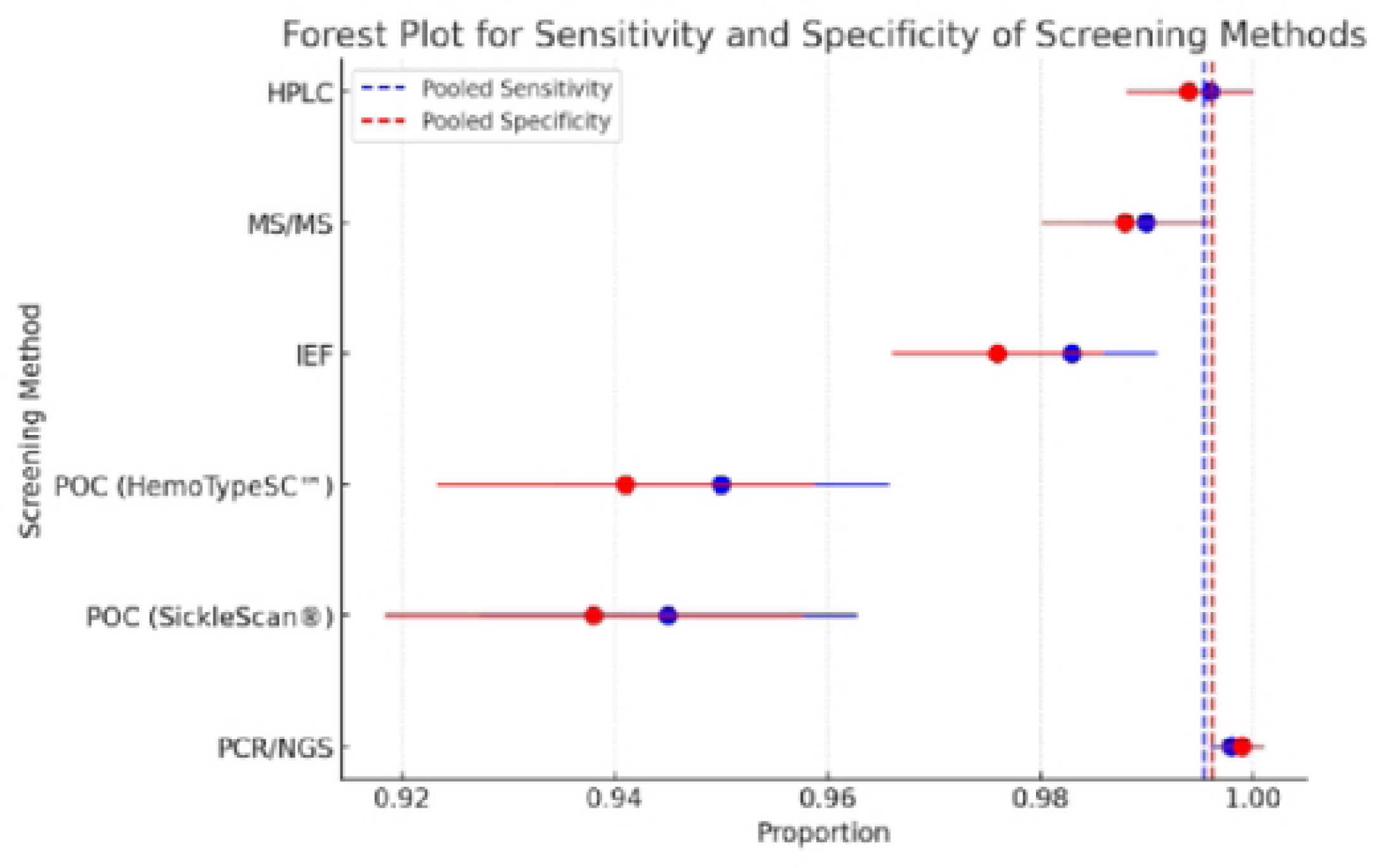
Forest Plot for sensitivity and specificity of screening methods.

To improve global consistency in sickle cell disease detection, the following recommendations are proposed:

- Standardize screening protocols, ensuring that the tests used are validated in multiple settings and populations.
- Optimize confirmatory methods, combining initial screening tests with molecular techniques in uncertain cases.
- Strengthening infrastructure in low-resource settings, facilitating access to more precise methods such as HPLC or PCR.
- Train healthcare personnel in test interpretation, minimizing errors in reading and classifying results.

## 4. Discussion

Sickle cell disease (SCD) is hereditary hemoglobinopathy that can cause severe complications from the first months of life. Early detection through neonatal screening allows for the timely implementation of preventive and therapeutic measures, significantly reducing the morbidity and mortality associated with this disease. Various screening methods have been developed and evaluated in terms of their diagnostic accuracy, including sensitivity, specificity, and heterogeneity across studies.

### Sensitivity in Neonatal Screening for SCD

Sensitivity and specificity are fundamental metrics in the evaluation of neonatal screening methods for sickle cell disease (SCD). Sensitivity refers to a test’s ability to correctly identify individuals with the disease (true positives), while specificity measures its ability to correctly classify those without the disease (true negatives) [16]. High values in both metrics are crucial in ensuring accurate and early diagnosis, minimizing false positives and false negatives, and optimizing healthcare interventions.

The pooled sensitivity of screening methods for SCD in our meta-analysis was 99.54% (95% CI: 99.38% - 99.71%), demonstrating that current screening techniques are highly effective at detecting affected newborns. High-Performance Liquid Chromatography (HPLC), Tandem Mass Spectrometry (MS/MS), and Polymerase Chain Reaction (PCR)/Next-Generation Sequencing (NGS) methods achieved the highest sensitivity, exceeding 99% in most studies [17,18].

A high sensitivity is essential in neonatal screening programs, as missing an affected infant could delay crucial interventions such as penicillin prophylaxis, vaccinations, and hydroxyurea therapy, which significantly reduce morbidity and mortality in children with SCD [19]. However, even small reductions in sensitivity can have clinical consequences, particularly in low-resource settings where opportunities for follow-up screening may be limited.

In contrast, Point-of-Care (POC) tests, such as HemoTypeSC™ and SickleScan®, demonstrated slightly lower sensitivity, ranging from 91.3% to 98.6%, depending on study conditions and sample handling [20]. While these values still indicate good diagnostic performance, POC tests may miss some affected newborns, particularly those with hemoglobin variants that are more challenging to detect.

### Specificity in Neonatal Screening for SCD

The pooled specificity in our analysis was 99.62% (95% CI: 99.44% - 99.79%), indicating that the evaluated screening methods are also highly effective at excluding unaffected individuals. Specificity is particularly important to minimize false positives, which can lead to unnecessary anxiety for parents, additional confirmatory testing, and increased healthcare costs [25].

Among laboratory-based methods, HPLC and MS/MS consistently reported specificities above 99%, reinforcing their gold standard status in neonatal screening. These tests effectively differentiate HbS from other hemoglobin variants (e.g., HbC, HbE, and thalassemias), reducing misclassification errors [21].

POC tests, while more accessible and affordable, exhibited lower specificity, ranging from 88.9% to 98.3% depending on the study. This increased false-positive rate is attributed to environmental factors, test variability, and operator-dependent errors, which may lead to unnecessary follow-up testing [22]. Despite this, POC methods remain invaluable in regions where laboratory-based screening is not available, as their rapid results enable early intervention.

### Balancing Sensitivity and Specificity in Screening Programs

In an ideal scenario, a perfect screening test would have both 100% sensitivity and specificity. However, in real-world applications, there is often a trade-off between the two metrics. Programs prioritizing sensitivity might accept some false positives to ensure that no affected newborns are missed. Conversely, those emphasizing specificity might reduce false positives at the risk of missing some cases, which could delay crucial medical interventions.

The decision on which screening method to implement should be considered:

- Healthcare infrastructure: Countries with strong laboratory capacity can rely on HPLC, MS/MS, or PCR-based confirmation, while low-resource regions may depend on POC screening with follow-up confirmatory testing [23].
- Cost-effectiveness: While laboratory-based methods offer superior accuracy, POC tests provide a lower-cost alternative with rapid results, making them suitable for mass screening programs [24].
- Population characteristics: Different regions have varying prevalence rates of hemoglobinopathies, which may influence the choice of screening strategies. In sub-Saharan Africa, where SCD prevalence is highest, sensitivity may be prioritized to ensure no affected newborn is missed [25].

Addressing Variability and Improving Screening Accuracy

Given the observed heterogeneity (I² Sensitivity: 93.99%, I² Specificity: 95.02%), improving the consistency of screening outcomes requires:

1. Standardization of testing protocols across different settings to reduce inter-study variability.
2. Improved training for healthcare professionals administering and interpreting POC tests to minimize user-dependent errors.
3. Hybrid screening models, integrating POC tests for initial detection with laboratory confirmation for high-risk cases, optimizing both cost and accuracy [26].
4. Further research into AI-based screening approaches, where machine learning models can improve the interpretation of screening results, particularly in low-resource settings [27].

### Diagnostic Accuracy of Screening Methods

Neonatal screening methods for SCD have generally demonstrated high diagnostic accuracy. Techniques such as high-performance liquid chromatography (HPLC), tandem mass spectrometry (MS/MS), and isoelectric focusing (IEF) are widely used due to their high sensitivity and specificity. For example, a report from the Galician Agency for Health Technology Assessment indicates that these methods can achieve sensitivity and specificity close to 100%, making them reliable tools for early disease detection [28].

Additionally, point-of-care (POC) tests, such as HemoTypeSC™ and SickleScan®, have been developed to facilitate screening in resource-limited settings. Although these methods offer advantages in terms of speed and ease of use, some studies have noted that their accuracy may be slightly lower compared to traditional laboratory techniques. For instance, the Spanish Society of Hematology and Hemotherapy (SEHH) guidelines emphasize that while these rapid tests are useful, their results should be confirmed using laboratory methods due to possible variations in accuracy [29].

### Heterogeneity Across Studies

Heterogeneity among studies is a critical factor when assessing the effectiveness of screening methods. The I² statistic is commonly used to quantify heterogeneity, indicating the percentage of variability in results that is due to differences between studies rather than chance. I² values above 75% are considered indicative of high heterogeneity [21].

In the context of neonatal screening for SCD, significant heterogeneity has been observed in sensitivity and specificity metrics reported in different studies. This variability may be attributed to several factors, including differences in screening protocols, sample quality, study populations, and laboratory conditions. For instance, a study conducted in the Balearic Islands highlighted the need to standardize screening methods to reduce variability in results and improve comparability across different regions [19].

### Clinical Implications and Recommendations

The high diagnostic accuracy of neonatal screening methods for SCD supports their implementation in public health programs. However, the observed heterogeneity among studies underscores the need to standardize screening protocols and ensure quality at all stages of the diagnostic process. The following recommendations are suggested:

1. Standardization of Protocols: Develop and adopt clinical guidelines that clearly define screening procedures, including sample collection, test selection, and result interpretation criteria.
2. Healthcare Staff Training: Ensure that healthcare professionals involved in screening are adequately trained in performing and interpreting screening tests, minimizing errors and ensuring diagnostic quality.
3. Quality Control Programs: Implement quality control measures in laboratories performing screening tests to ensure consistency and reliability of results.
4. Continuous Evaluation: Conduct periodic assessments of screening programs to identify areas for improvement and adapt strategies based on the specific needs of the population.

## 5. Conclusion

Although neonatal screening methods for SCD have demonstrated high diagnostic accuracy, addressing heterogeneity across studies through the standardization of protocols and the implementation of quality control measures is essential. Ensuring universal access to early and precise diagnosis will facilitate timely interventions that improve long-term health outcomes for newborns affected by sickle cell disease.

NBS for SCD has evolved significantly, with advancements in biochemical, molecular, and AI-driven methods enhancing early detection. While HPLC and MS/MS remain gold standards, point-of-care and genomic approaches hold promise for expanding access, particularly in underserved regions. Future research should focus on optimizing cost-effective screening strategies and integrating AI for more efficient diagnostics.

## Data Availability

The data that support the findings of this study are available on request from the corresponding author. The data are not publicly available due to privacy or ethical restrictions.

